# Stability of SARS-CoV-2 on Critical Personal Protective Equipment

**DOI:** 10.1101/2020.06.11.20128884

**Authors:** Samantha B Kasloff, James E Strong, Duane Funk, Todd Cutts

**Affiliations:** National Microbiology Laboratory, Public Health Agency of Canada, 1015 Arlington Street, Winnipeg, Manitoba, R3E 3R2, Canada; Department of Pediatrics & Child Health, College of Medicine, Faculty of Health Sciences, University of Manitoba, Winnipeg, Manitoba, Canada; Department of Infectious Diseases and Medical Microbiology, College of Medicine, Faculty of Health Sciences, University of Manitoba, Winnipeg, Manitoba; Department of Anaesthesia and Medicine, College of Medicine, Faculty of Health Sciences, University of Manitoba, Winnipeg, Manitoba, Canada

**Author notes:** These authors contributed equally to this work (Samantha B Kasloff, Todd Cutts).

## Abstract

The spread of COVID-19 in healthcare settings is concerning, with healthcare workers representing a disproportionately high percentage of confirmed cases. Although SARS-CoV-2 virus has been found to persist on surfaces for a number of days, the extent and duration of fomites as a mode of transmission, particularly in healthcare settings, has not been fully characterized. To shed light on this critical matter, the present study provides the first comprehensive assessment of SARS-CoV-2 stability on experimentally contaminated personal protective equipment (PPE) widely used by healthcare workers and the general public. Persistence of viable virus was monitored over 21 days on eight different materials, including nitrile medical examination gloves, reinforced chemical resistant gloves, N-95 and N-100 particulate respirator masks, Tyvek®, plastic, cotton, and stainless steel. Unlike previous reports, viable SARS-CoV-2 in the presence of a soil load persisted for up to 21 days on experimentally inoculated PPE, including materials from filtering facepiece respirators (N-95 and N-100 masks) and a plastic visor. Conversely, when applied to 100% cotton fabric, the virus underwent rapid degradation and became undetectable in less than 24 hours. These findings underline the importance of appropriate handling of contaminated PPE during and following use in high-risk settings and provide interesting insight into the potential utility of cotton, including cotton masks, in limiting COVID-19 transmission.

## Introduction

Since its emergence in China at the end of 2019, the unprecedented spread of SARS-CoV-2 has lead to the declaration of a pandemic by the World Health Organization. The role of healthcare settings in the spread of this virus is of particular concern, as healthcare workers have represented a disproportionately high number of confirmed cases in the first months of the COVID-19 pandemic (COVID-19 Editorial, 2020; Anelli *et al*, 2020; Time.com; CEBM 2020; CBC 2020; LA Times). Similar trends were noted in the 2003 SARS outbreak, where healthcare workers accounted for approximately 20% of cases (Chan-Yeung, 2004).

Among the many questions yet to be adequately addressed about the modes of transmission of COVID-19 are those related to fomite spread and environmental contamination. In healthcare settings in particular, numerous surfaces ranging from door handles to disposable gowns and gloves may become contaminated by droplets of infectious secretions and shed by infected persons. Further, global shortages in personal protective equipment (PPE) may result in items intended for single use being reused or worn for longer periods than recommended (Ranney, 2020). This in turn may increase the opportunity for viral contamination and subsequent spread. An understanding of the environmental stability and persistence of SARS-CoV-2 on contaminated PPE may have profound impacts on the handling of both reusable and single-use items during use after wear.

The present study aimed to determine the stability of SARS-CoV-2 on experimentally inoculated surfaces of PPE widely used by healthcare workers and the members of the general population. Our findings underline the importance of appropriate handling of contaminated PPE after its use and provide interesting insight into the potential utility of cotton masks in limiting COVID-19 transmission.

## Materials and Methods

### Cell culture and virus

Passage 3 stocks of SARS-CoV-2 (*hCoV-19/Canada/ON-VIDO-01/2020, GISAID accession# EPI_ISL_425177*) were prepared in Vero E6 cells as previously described (Cook et al, 2015). Endpoint titration of stock virus and all experimental eluates were calculated via the 50% Tissue Culture Infectious dose assay (TCID_50_) on Vero E6 cells in a 96 well plate format using the method of Reed and Muench (1938). As SARS-CoV-2 is classified as a Risk Group 3 pathogen, all experimental procedures took place in a high containment laboratory at the National Microbiology Laboratory in Winnipeg, Canada.

#### Test Surfaces

Eight materials, representing a range of environmental surfaces commonly encountered in healthcare facilities with a particular focus on personal protective equipment were included in this study. These included nitrile medical examination gloves, reinforced chemical resistant gloves, N-95 and N-100 particulate respirator masks, Tyvek® coveralls, plastic from face shields, heavy cotton, and stainless steel (Table 1).

**Table 1.** Comprehensive descriptions of materials used for assessment of SARS-CoV-2 environmental persistence.

| Test Material | Description | Manufacturer/Source | Porosity |
| --- | --- | --- | --- |
| Stainless Steel | Stainless Steel (10mm, AISI 430) | Klassen Manufacturing, Winnipeg Canada | Non-porous |
| Plastic face shield | Plastic face shield from PAPR Hood Assembly (BE-10L) | 3M™, Saint Paul, Minnesota | Non-porous |
| Nitrile gloves | Nitrile Gloves (KC55081) | Kimberly-Clark Professional™, Irving, Texas | Non-porous |
| Chemical gloves | Chemical resistant, reinforced nitrile gloves (AlphaTec® 39-124) | Ansell, Melbourne, AUS | Non-porous |
| Tyvek | Tyvek® 400 Coveralls (TY125SWH) | DuPont™, Wilmington, Delaware | Porous |
| Mask 1 | N95 Particulate Filter Respirator and Surgical Mask (KC46727) | Kimberly-Clark Professional™, Irving, Texas | Porous |
| Mask 2 | N-100 Particulate Respirator (3M8233) | 3M™, Saint Paul, Minnesota | Porous |
| Cotton | Heavy Cotton T Shirt | Fruit of the Loom, Bowling Green, Kentucky | Porous |

Pre-cut, 1cm^2^ stainless steel coupons were obtained directly from the manufacturer and sterilized as previously described (Springthorpe & Sattar, 2003). Coupons of all other experimental materials were cut onsite to 1.4 cm^2^ and sterilized by 2 Mrads of gamma radiation prior to use.

### Experimental Inoculation and Sampling

Experimental inoculum consisted of stock virus (titre = 7.88 LogTCID_50_/mL) in a tripartite soil load containing mucin (Sigma #M3895), bovine serum albumin (Sigma #A1933) and tryptone (Sigma #T7293) to represent the organic components of typical virus-containing fluids shed by infected individuals (ASTM 2002; Sattar et al 2003). Ten microliters of the resulting virus suspension was added via positive displacement pipette to the center of each coupon and left to air dry in a biosafety cabinet for 1 hour. Once dry, coupons were placed in a vented plastic container and stored in a closed cabinet at ambient temperature for the duration of the study. Daily recordings of temperature and humidity at 12-hour intervals were monitored with a Professional Thermo-Hygrometer with data logger (TFA Dostmann Product#30.3039.IT).

Three technical replicates (N=3) of each test material were sampled at the following time points post inoculation: T=1hr (point of drying), 4hrs, and at 1, 2, 3, 4, 7, 14 and 21 days. Virus was recovered by elution in 1 mL of culture medium (DMEM + 2% FBS + 1% Pen-Strep) with rigorous and repeated pipetting. Eluates were immediately 10-fold serially diluted for endpoint titration in Vero E6 cells, using three biological replicates per dilution. Remaining volumes of eluates were stored at -80°C for potential re-processing if required. Samples resulting in low-level cytopathic effect (i.e., in only a single well at neat dilutions) underwent additional sub-passage to confirm the presence of viable virus. Additionally, on the first instance where all technical replicates of a given sample material resulted as negative, neat dilutions of all three biological replicates underwent additional sub-passage to ensure capture of potentially low-level virus not visually detected on the first passage. At the completion of the study, the full remaining volume from each day 21 eluate was added to an individual well of a 6-well plate of Vero E6 cells and monitored for cytopathic effect to ensure even a single viable virus particle was captured.

### Statistical Analyses

Graphical representations of TCID_50_ results, including averages and standard deviations, were performed using GraphPad Prism (version 7) software. Technical replicates with no recoverable virus were assigned a value of zero for the purposes of these calculations.

## Results

The environmental persistence of SARS-CoV-2 dried onto surfaces of commonly used PPE and surfaces found in healthcare settings over a period of 21 days revealed dramatic differences in virus stability. By 4 hours post-deposition on cotton, infectious SARS-CoV-2 virus was drastically diminished, and no longer recoverable by 24 hours. Conversely, while reduced almost to the limit of detection, SARS-CoV-2 remained viable for 21 days when dried on plastic and particulate respirator procedural masks. Real-time monitoring of ambient conditions revealed a steady temperature near 20°C with 35-40% relative humidity (**Figure 1**).

**Figure 1.**
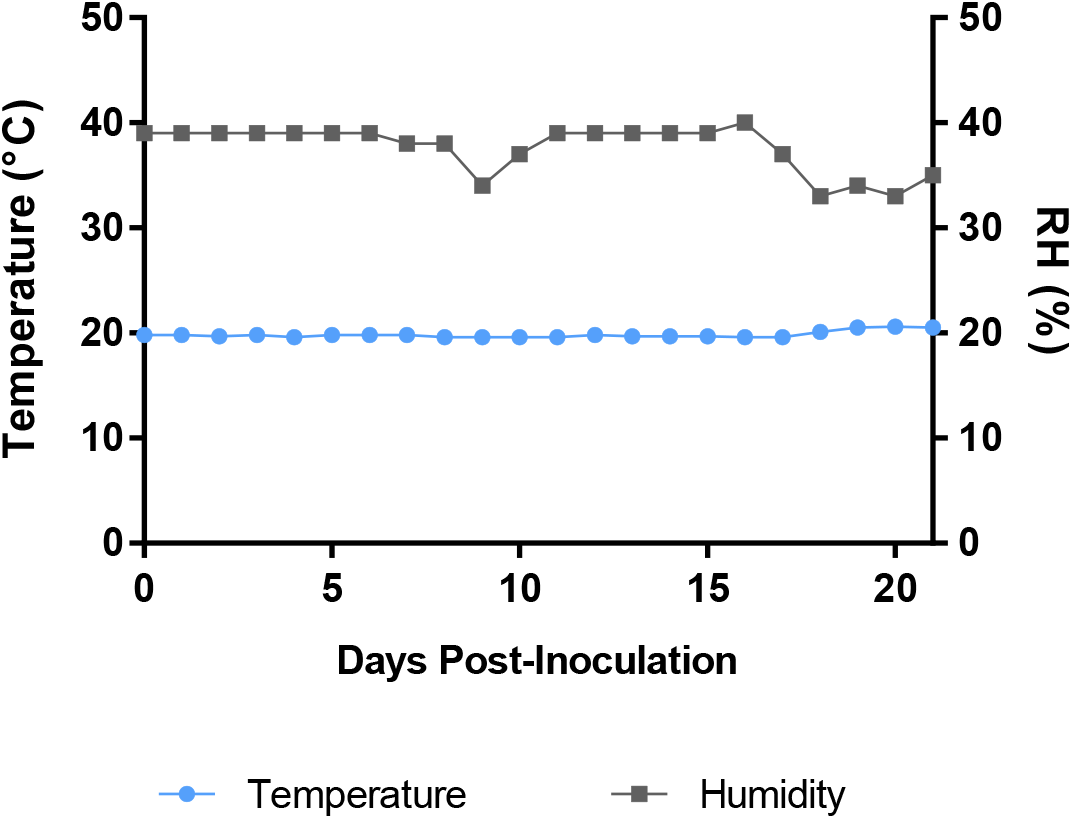
Ambient conditions recorded for the duration of the experiment. Temperature (°C) and relative humidity (%RH) readings were logged at 12-hour intervals using a Professional Thermo-Hygrometer with data logger.

### Persistence of SARS-CoV-2 on Non-Porous surfaces

When a dried onto non-porous surfaces in an organic soil load SARS-CoV-2 showed sustained persistence for a prolonged period of time. Viable SARS-CoV-2 was recovered after 21 days on plastic, 14 days on stainless steel, 7 days on nitrile gloves and 4 days on chemical resistant gloves, though at significantly reduced levels compared to the initial inoculum (**Table 2, Figure 2**).

**Table 2.** Table 2. Recovery of viable SARS-CoV-2 on experimentally inoculated environmental surfaces (Log10 TCID_50_/mL)

| Sampling Time | Stainless steel |  | Tyvek |  | Plastic |  | Nitrile gloves |  | Chemical gloves |  | Mask 1 |  | Mask 2 |  | Cotton |  |
| --- | --- | --- | --- | --- | --- | --- | --- | --- | --- | --- | --- | --- | --- | --- | --- | --- |
|  | Mean<sup>a</sup> | +/- SD | Mean | +/- SD | Mean | +/- SD | Mean | +/- SD | Mean | +/- SD | Mean | +/- SD | Mean | +/- SD | Mean | +/- SD |
| 1 hr | 5.58 | 0.14 | 5.50 | 0.00 | 5.75 | 0.66 | 5.92 | 0.29 | 3.42 | 2.01 | 5.75 | 0.50 | 5.75 | 0.25 | 1.42 | 0.14 |
| 4 hrs | 5.25 | 0.50 | 5.17 | 0.52 | 5.92 | 0.63 | 5.67 | 0.52 | 2.25 | 1.30 | 5.50 | 0.25 | 5.17 | 0.14 | 1.08 | 0.95 |
| 1 day | 4.39 | 0.13 | 4.50 | 0.43 | 4.50 | 0.25 | 3.67 | 0.14 | 1.58 | 0.14 | 4.67 | 0.14 | 4.58 | 0.80 | - |  |
| 2 days | 3.58 | 0.14 | 3.58 | 0.14 | 3.75 | 0.43 | 3.00 | 0.66 | 1.92 | 0.29 | 3.75 | 0.00 | 3.58 | 0.38 | - |  |
| 3 days | 3.00 | 0.00 | 3.33 | 0.14 | 3.42 | 0.14 | 2.67 | 0.14 | 1.58 | 0.14 | 3.50 | 0.25 | 3.33 | 0.29 | - |  |
| 4 days | 3.33 | 0.14 | 3.00 | 0.25 | 3.42 | 0.14 | 3.25 | 1.09 | 1.75 | 0.43 | 3.75 | 0.43 | 3.33 | 0.29 | - |  |
| 7 days | 2.42 | 0.29 | 2.00 | 0.25 | 2.00 | 0.43 | 1.50 | 0.25 | - |  | 2.33 | 0.38 | 2.33 | 0.38 | - |  |
| 14 days | 0.75 | 0.66 | 0.67 | 0.63 | 1.01 | 0.24 | - |  | - |  | 1.26 | 0.42 | 1.17 | 0.37 | - |  |
| 21 days | - |  | - |  | 0.26* | 0.44 | - |  | - |  | 0.83* | 0.76 | 0.26* | 0.44 | - |  |

<sup>a</sup> Mean TCID<sub>50</sub> values calculated from three independent technical replicates, each with triplicate biological replicates per dilution. Limit of detection was Log(0.767) TCID<sub>50</sub>/mL
( - ) = no CPE detected in any biological replicate at any dilution of eluted material.
\* presence of viable virus confirmed upon subpassage

**Figure 2.**
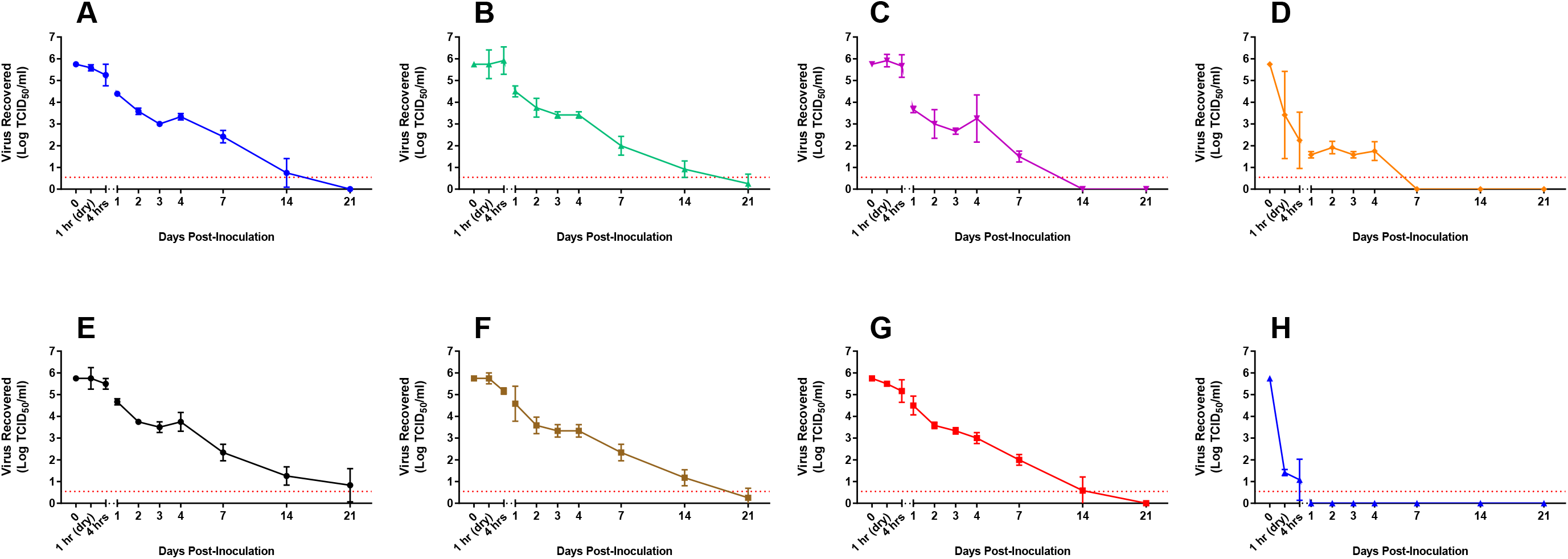
Persistence of viable SARS-CoV-2 on experimentally inoculated surfaces. Materials from eight experimental surfaces, including A) stainless steel, B) plastic, C) nitrile gloves, D) chemical gloves, E) N95 mask, F) N100 mask, G) Tyvek, and H) cotton were inoculated with 10 μl of SARS-CoV-2 (titre = 7.88 LogTCID_50_/mL) in a tripartite soil load and sampled at predetermined intervals over a 21 day time-course. All materials were sampled in triplicate (n=3) at each experimental time point by elution in 1 mL culture medium, with viable virus quantified by end-point titration in Vero E6 cells. Results represent mean ± SD log TCID_50_ values of recovered virus from three technical replicates. Red dashed lines indicate limits of quantification for the TCID_50_ assay.

Persistence data on stainless steel showed a decrease in viral titre from 5.74 to 4.39 log TICD_50_/ml after a 24 hours, representing a >95% decrease in viable virus. Sampling over a 21-day time course revealed that SARS-CoV-2 remained viable on stainless steel for up to 14 days at ambient conditions, though final titers of 0.7 Log TCID_50_/ml were exponentially lower than starting inoculum (**Figure 2A**). On plastic, a similar reduction in viable virus was observed by 24 hours of drying compared to initial inoculum (from 5.74 to 4.5 log TCID_50_/ml). However, virus dried on this surface remained viable through the final sampling point at 21 days post-inoculation (**Figure 2B**).

Two glove types, representing those used in clinical and potential field hospital situations, were also included for analysis. With nitrile gloves, no decrease in viable virus was observed after four hours of drying, though by 24 hours a 2-log reduction in viral titer was recorded. At 7 days post-inoculation, low levels of viable virus (mean 1.50 log TCID_50_/ml) remained on all three technical replicates (**Figure 2C**). Conversely, virus applied to the chemical protective gloves showed a 2-log reduction in viability after only a single hour of drying, and a nearly 4-log reduction to 1.58 log TCID_50_/ml by 24 hours (**Figure 2D**).

### Persistence of SARS-CoV-2 on Porous Materials

Though reduced to almost undetectable levels, viable SARS-CoV-2 could be recovered from inoculated N-95 (Mask 1) and N-100 (Mask 2) surface materials for up to 21 days. Decay patterns of virus viability were comparable on the two masks; both showing a nearly 1 log decrease from 24-48 hours, stable titres between 2-4 days, and a steady decline from days 7 through 21 (**Figure 2E&F**). On Tyvek, infectious SARS-CoV-2 persisted up to 14 days; again, reduced to very low levels compared to the starting inoculum. Persistence patterns on this material (***Figure 2G***) most closely resembled those for stainless steel.

Of all the materials tested, cotton provided the lowest environmental stability to SARS-CoV-2. After only a single hour of drying, over 4 logs of viable virus were lost, representing a 99.995% decrease from input inoculum. After four hours of drying, a further decrease was recorded and only two of three technical replicates produced viable virus upon elution. By the 24 hour time point and at all subsequent sampling days, no detectable SARS-CoV-2 remained on cotton (**Figure 2H**).

## Discussion

The COVID-19 pandemic has led to unprecedented burdens on healthcare facilities worldwide. While respiratory droplets are considered an important mode of transmission, the role of fomites in the spread of SARS-CoV-2, as suggested for SARS-CoV-1 (Xiao et al), is also suspected (Ong et al). Our work represents the first comprehensive characterization of SARS-CoV-2 persistence on PPE materials and inanimate surfaces typical of healthcare facilities.

Viable SARS-CoV-2 was recovered after 21 days on plastic, 14 days on stainless steel, 7 days on nitrile gloves and 4 days on chemical resistant gloves. Though reduced from baseline, significant quantities of viable SARS-CoV-2 could be recovered from inoculated N-95 and N-100 masks at 14 days. When dried onto Tyvek, infectious SARS-CoV-2 persisted up to 14 days. Of all the materials tested, cotton provided the lowest environmental stability to SARS-CoV-2. After only a single hour of drying, over 4 logs of viable virus were lost, representing a 99.995% decrease from input inoculum. These results have direct relevance to IPC practices, laundering and waste handling protocols in healthcare settings.

The global shortage of N-95 masks has led healthcare centers worldwide to extend the usage of these masks despite them being designed for single use (Ranney, 2020). Our results demonstrate that this decision, in the absence of a decontamination strategy, may result in persistence of SARS-CoV-2 on the mask. Persistence of other viruses, such as influenza, Ebolavirus, and other coronaviruses, has been reported on experimentally-inoculated respirators (Coulliette et al, 2013; Cook et al 2015, Casanova et al 2010b). While a number of decontamination methodologies show promise for the re-use of N-95s (Fischer et al 2020; Kumar et al 2020), our results suggest that careful attention must be paid to their collection criteria and provision of appropriate risk-based PPE to staff involved in sorting and packaging of these masks prior to decontamination.

Our finding that SARS-CoV-2 can remain viable for up to two weeks at room temperature on Tyvek is novel and significant. Tyvek garments have been widely adopted in high-risk healthcare, field, and laboratory settings due to unique tensile properties combined with imperviousness to infectious agents. Viral persistence on Tyvek, combined with our demonstration of viral persistence on plastic has implications for the re-use of Powered Air Purifying Respirator hoods (PAPRs) in health care settings. Disinfection of these devices has proven to be challenging (Lawrence et al 2017). These results highlight the need for risk-based decision-making and implementation of appropriate decontamination protocols where re-use of these critical PPE components is practiced.

The extremely limited viability of SARS-CoV-2 on cotton aligns with survival results of other enveloped viruses on cotton (Cook et al, 2015; Tiwari et al, 2006; Bean et al 1982). Similar to our results with SARS-CoV-2, the viability of SARS-CoV-1 when applied at 10^5^ TCID_50_ /ml was limited to 1 hour on a cotton gown compared to 24 hours on a disposable gown (Lai et al 2005).

The use of non-medical masks to limit the spread of transmissible respiratory diseases, including COVID-19, has been contentious. While mounting evidence suggests that simple cloth (cotton) masks would be effective in limiting SARS-CoV-2 (Howard et al, 2020), others have questioned the efficacy of manufactured or home-made cloth masks to effectively prevent release of viral particles from infected individuals (Bae et al 2020, Davies et al 2013). Our observations in the present study are not intended for inference on the ability of cotton masks to prevent aerosol transmission of SARS-CoV-2, nor do they address filtration efficacy of cotton against the virus. However, the fact that virus viability is rapidly reduced on cotton exposure has implications for droplet transmission for both the wearer and nearby contacts, and complements the growing support of widespread cotton mask use. Further, these results suggest that the use of cotton-based fabrics in healthcare settings may present a lower risk during handling for subsequent decontamination and reuse.

The survival of SARS-Cov-2 on environmental surfaces has been described in two other studies (Chin et al Lancet Microbe 2020; van Doremalen et al NEJM 2020). Our results are in agreement with those reported, however viral persistence was significantly prolonged under our experimental conditions. These discrepancies are likely due to a combination of different ambient conditions, inoculum titres, and readout assay sensitivity.

The work of Chin and colleagues, using a 5 μl droplet of high titre stock virus (∼7.8 log TCID_50_ units/mL), most closely resembled our inoculation conditions. The reported 65% RH in their study compared to ∼40% RH under our experimental conditions is likely a significant contributor to the reduction in viability over time. The detrimental effects of high relative humidity on survival of other coronaviruses, including MERS and SARS-CoV-1, has been demonstrated (van Doremalen et al 2013; Chan et al., 2011; Casanova et al 2010a). Nevertheless, the availability of data specific to diverse environmental conditions is of importance to capture the realities at different geographical locations.

In the report by Van Doremalen et al., viable SARS-CoV-2 persisted up to only 3 days on stainless steel and plastic. While the reported ambient conditions more closely represented ours, the titre of infectious SARS-CoV-2 applied to test surfaces was approximately 2 logs lower than that applied in our study. The protective effect of high titre inoculum on environmental persistence has been previously shown for SARS-CoV-1 (Lai et al 2005) and therefor the extended survival observed under our experimental design is not surprising.

An additional factor differentiating the present work from other studies is the matrix in which inocula were prepared. Contamination in a healthcare setting can occur through droplets from an array of infectious biological fluids, therefore a standardized tripartite soil load (ASTM, 2002) was included in our experimental inoculum. The added stability provided by matrices to viruses under drying conditions has been demonstrated for many viruses, including SARS-CoV-1 (Rabenau et al, 2005; Sattar et al 2003, Terpstra et al 2007; Krug et al 2011). As neither of the two recently published studies mention the use of a soil load for testing environmental stability, this likely accounts for the extended viral persistence observed in our experimental setting.

The decision to use a high titre inoculum in our study was to represent a worst-case scenario of SARS-CoV-2 persistence on a contaminated surface. Shedding of viral RNA from COVID-19 patients varies widely based on sample type, time post-symptom onset, severity of disease, and on an individual basis (Zou et al 2020, Pan et al 2020). Clinical data from mild and severe cases has revealed a range of data, with median initial viral loads of 5.11 and 6.17 log_10_ copies of viral RNA per ml in mild and severe cases, respectively (To et al 2020). In a critically ill patient, loads of up to 1.34 × 10^11^ copies per mL were detected (Pan et al, 2020). Data on infectious virus shed in excreta from acutely ill COVID-19 patients is lacking. However, in a recently published SARS-CoV-2 pathogenesis study, peak viral loads of >6 log_10_ TCID_50_/mL, corresponding to 11 log_10_ TCID_50_ RNA copies per mL were observed in the golden hamster model (Sia et al 2020). Taking these data into consideration, our experimental model would more closely demonstrate environmental contamination from a severe COVID-19 case.

A potential limitation of this study lies in testing being carried out in a high containment laboratory, where 12 air exchanges per hour may represent a high air flow not necessarily representative of most health facilities, homes and other environments. Additionally, the initial hour of drying time under a biosafety cabinet would have accelerated the drying effect and may have shortened the amount of time to killing of the virus. However, the revealed persistence of up to 21 days was significant and may be the best-case scenario associated with high airflow conditions that can occur in some health care environments.

Additionally, the implications of these results and those of others (Chin, Van Doremalen) on the duration of potential fomite transmission of SARS-CoV-2 should be interpreted with caution due to the sampling methodology utilized. Viral recovery was achieved by material saturation in media, enabling recovery of extremely low levels of remaining virus. Had we employed a swabbing method to more closely mimic a casual contact scenario, a decrease in sensitivity would be expected and thus duration of viable virus detection may have been reduced. As a result, the data generated represent a worst-case scenario for potential exposure through these contaminated surfaces, though the risk of fomite transmission through casual contact can only be inferred by the presence of infectious virus at a given time point.

Our findings suggest that SARS-CoV-2 can remain infectious on contaminated PPE for extended periods under ambient environmental conditions. Conversely, they demonstrate that stability is highly reduced on cotton even within hours of contamination, and encourage the use of 100% cotton in infection prevention and control practices in healthcare settings and general use. These results underline the importance of proper handling of personal protective equipment during and following use in high-risk settings to minimize the likelihood of fomite transmission, and may assist centers in developing guidelines and protocols relating to decontamination and reuse of PPE in short supply.

## Data Availability

Data and associated protocols can be made available to readers without undue qualifications in material transfer agreements.

## Acknowledgments

We kindly thank Dr. Yohannes Berhane for his insightful review of this manuscript. The authors declare no potential conflicts of interest.

